# Parkinson’s disease impairs cortical sensori-motor decision-making cascades

**DOI:** 10.1101/2022.10.26.22281571

**Authors:** A. Tomassini, T. E. Cope, J. Zhang, J. B. Rowe

**Affiliations:** MRC Cognition and Brain Sciences Unit, University of Cambridge. 15 Chaucer Road, Cambridge, CB2 7EF, UK; Department of Clinical Neurosciences, University of Cambridge. Herchel Smith Building, Forvie Site Robinson Way, Cambridge Biomedical Campus, Cambridge, CB2 0SZ, UK; Cambridge University Hospitals NHS Trust, Cambridge, CB2 0QQ, UK; Department of Computer Science, Swansea University, Swansea, SA18EN, UK

**Keywords:** Parkinson’s disease, accumulator, uncertainty, visuomotor, decision-making

## Abstract

The transformation from perception to action requires a set of decisions about the nature of the percept, identification and selection of response options, and execution of the appropriate motor response. The unfolding of such decisions is mediated by distributed representations of the decision variables - evidence and intentions – that are represented through oscillatory activity across the cortex. Here we combine magneto-electroencephalography and linear ballistic accumulator models of decision-making to reveal the impact of Parkinson’s disease during the selection and execution of action. We used a visuomotor task in which we independently manipulated uncertainty in sensory and action domains. A generative accumulator model was optimized to single-trial neurophysiological correlates of human behaviour, mapping the cortical oscillatory signatures of decision-making, and relating these to separate processes accumulating sensory evidence and selecting a motor action. We confirmed the role of widespread beta oscillatory activity in shaping the feed-forward cascade of evidence accumulation from resolution of sensory inputs to selection of appropriate responses. By contrasting the spatiotemporal dynamics of evidence accumulation in age-matched healthy controls and people with Parkinson’s disease, we identified disruption of the beta-mediated cascade of evidence accumulation as the hallmark of atypical decision-making in Parkinson’s disease. In frontal cortical regions, there was inefficient processing and transfer of perceptual information. Our findings emphasize the intimate connection between abnormal visuomotor function and pathological oscillatory activity in neurodegenerative disease. We propose that disruption of the oscillatory mechanisms governing fast and precise information exchanges between the sensory and motor systems contributes to behavioural changes in people with Parkinson’s disease.

## Introduction

Parkinson’s disease is a neurodegenerative disorder characterised by the motor features of tremor, rigidity, bradykinesia and postural instability, associated with dopaminergic deficiency in the basal ganglia. However, cognitive and behavioural changes are common and early aspects of the disease, even before clinically manifest cognitive impairment and dementia. Impairments include processing speed, simple perceptual and complex behavioural decisions^1,2^ attention and selection of actions^3^, and inhibitory control^4^, which together underlie a wide range of executive function deficits^5^. The cognitive processes affected by Parkinson’s disease are not necessarily restricted to those mediated by neurons in the basal ganglia foci of early pathology, but may include widely distributed cortical networks.

The cognitive processes underlying the deficits in Parkinson’s disease can be conceived as a set of neuronal decisions, linking perception of environmental cues to the selection and execution of appropriate actions. Even apparently “simple” decisions unfold via sequential overlapping processes^6^, establishing a cascade in which the accumulation of evidence for perceptual decisions informs the accumulation of evidence (or ‘intentions’) for selection of a motor response^7–11^. These perceptual and motor decisions are not fully separated in time or space. A spatiotemporal overlap in the decisions enables an optimal compromise between robust but slow serial processes (i.e., action selection waits for the full resolution of the perceptual deliberation) and fast but error-prone parallel processes (i.e., synchronous action selection and perceptual decisions). Departures from optimal spatiotemporal overlap in the cascade of visuomotor decisions may bias towards slow or inaccurate responses.

Spatially distributed processing exploits neural oscillations for effective communication between regions^12^. Large-scale oscillatory activity at beta (∼13-30Hz) and more localized activity at gamma (∼30-90Hz) frequency ranges have been particularly associated with feedback and feedforward information transfer respectively^13–15^. Changes in spectral power and disruption of functional brain network organization occur in Parkinson’s disease. For example, exaggerated oscillations at beta frequency are a signature of pathology in the basal ganglia and frontoparietal network in Parkinson’s disease patients^16,17^ (for a review see Schnitzler, 2005^18^).

Dopaminergic deficiency in Parkinson’s disease may contribute to the generation of aberrant oscillatory activity in both the beta and gamma ranges^19^. A feature of pathological oscillations in Parkinson’s disease is the reduced power modulation elicited by changes in environmental and cognitive demands. Such inflexibility may contribute to cognitive changes in Parkinson’s disease^20^. This mirrors normative accounts of decision-making. For example, sequential sampling models of behaviour in which deficits are associated with inflexibility of the latent cognitive decision-processes estimated from patients’ performance on experimental tasks^2,21,22^. Distributed frontoparietal networks are involved in the selection and accumulation of evidence to reach decisions in non-human primates^10^ and humans^23,24^. Abnormalities in these circuits arise early in Parkinson’s disease. Their functional significance is revealed by integrating neurophysiology (EEG and/or MEG) with models of behaviour in which latent cognitive processes cannot be observed directly^25^.

Here we adopted Linear Ballistic Accumulator models to identify latent components of cognitive processes^26^ (LBA). The LBA posits separate accumulator processes, accruing evidence for alternatives in a race to reach the decision boundary. The fastest accumulator resolves the decision. In addition to time spent in evidence accumulation, there are *non-decision processes* before and after. The non-decision time includes sensory encoding and motor execution processes. These are not typically distinguished in behavioural modelling^27–29^, but it is possible to do so^25^. Decomposing the non-decision time enables one to examine the spatial distribution of the latency to accumulation^30^. This in turn enables the separation of the effect of spatially inhomogeneous disease on perceptual encoding *versus* noise of accumulators^2,21,30,31^. The transformation of perceptual decisions into action selection across the dorsal stream may be especially susceptible to the dopaminergic effect of Parkinson’s disease^10,23,32^. We predict that the effect of Parkinson’s disease on the spatiotemporal cascade of evidence accumulation would not only be seen within frontostriatal circuits, but also posterior cortex in receipt of frequency-bound top-down signalling from the frontal cortex.

To test this prediction, we used electro-magnetoencephalography (MEEG) in people with Parkinson’s disease, without dementia, on their usual medication. In the visuomotor decision task, noisy visual stimuli indicate one or more permitted manual response options (Figure 1A). We independently manipulated perceptual uncertainty and the number of permitted actions ^15^. This separated variance due to the neural signatures of perceptual decisions (e.g., *which options are available?*) from action decisions (e.g., *which option do I choose?*), without *a priori* spatial bias to “sensory” or “motor” cortical regions. The spatio-temporal pattern of decision onsets was identified by optimizing the division of the non-decision time to before *versus* after the accumulation period, according to trial-specific profiling of the induced power^15^.

**Figure 1.**
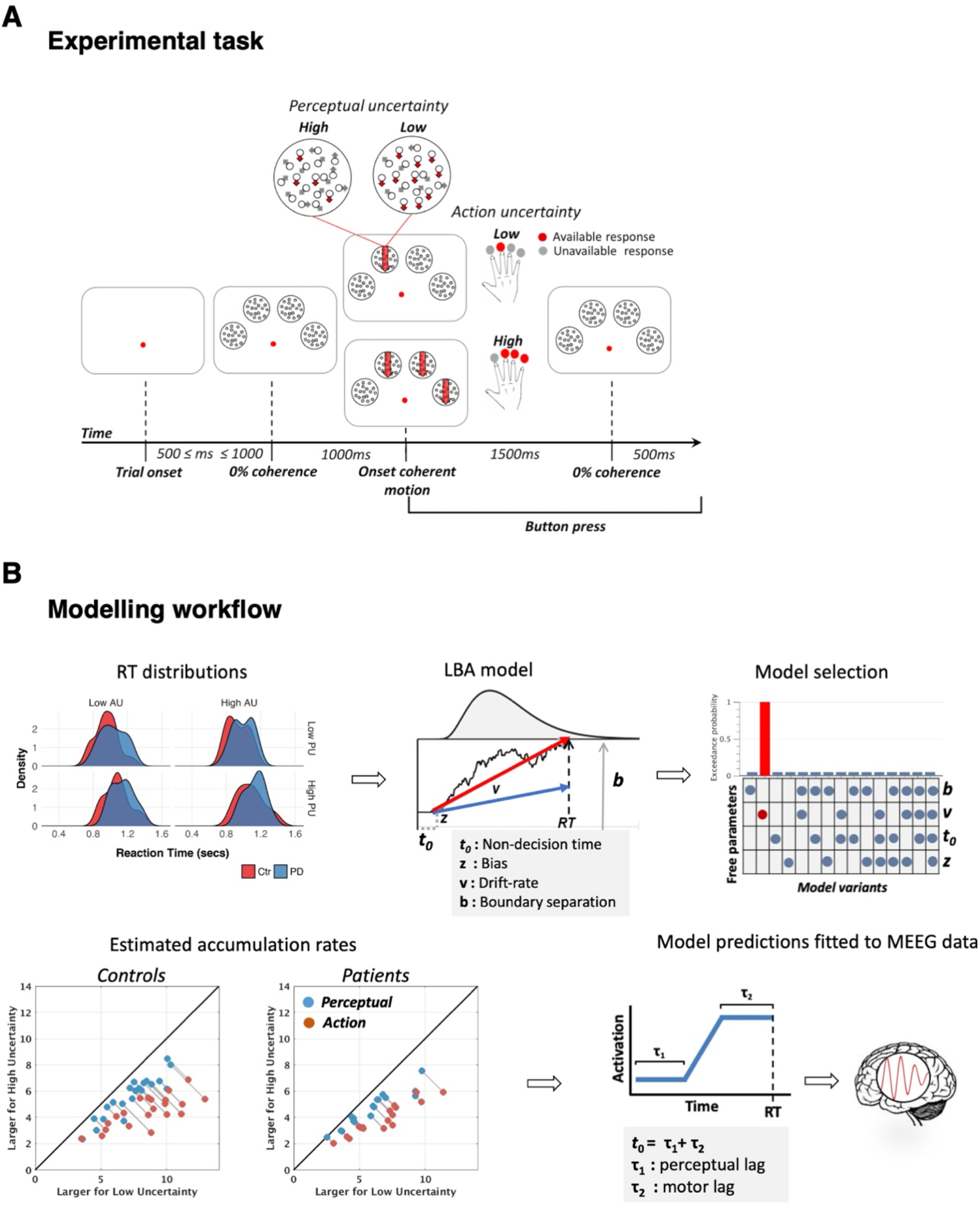
Experimental task and modelling procedure. **(A)** Participants pressed the button corresponding to the coherent stimulus (red downward arrow). When there was more than one coherent stimulus, they selected one response and pressed the corresponding button. Perceptual uncertainty was manipulated by changing the coherence of dot motion (i.e., by changing the motion strength), whereas action uncertainty was manipulated by changing the number of available options (i.e., the number of coherent stimuli to choose from). Perceptual and action uncertainty varied across trials in a 2 by 2 factorial design. **(B)** Reaction times from each participant (upper left panel) were modelled using a Linear Ballistic Accumulator (LBA) model (upper middle panel). Noisy evidence accumulates over time at a rate *v* up to a decision bound *b*. The fastest accumulator (thick red arrow) determines the choice. Non-decision time linked to sensorial and motor processes (*t0*) sums to the evidence accumulation time to produce reaction-times. The model best accounting for the behavioural data was selected with Bayesian model comparison (upper right panel): changes in the sole drift-rate best accounted for the behavioural data. For both controls and patients, the model predicted faster accumulation of decision evidence when uncertainty is low for both action and perceptual uncertainty (lower left panel; grey lines connect data points from each participant). Neural activity simulated with the winning model was fitted to the power envelope of the MEEG signal in a trial-by-trial fashion to identify the latencies of accumulators of decision evidence. Non decision time (*t*_*0*_) was decomposed into pre-accumulation (τ_1_) and post-accumulation (τ_2_) time reflecting perceptual and motor processes, respectively (lower right panel).

We predicted that people with Parkinson’s disease have shorter sensorial processing latency relative to controls^33,34^ and differential spatial gradients in the onset of frequency-specific accumulation across the dorsal stream. Whereas controls modulate the latent cognitive processes in response to the different levels of uncertainty (indexed by the LBA modelling), we predicted relative insensitivity to task modulation of uncertainty in people with Parkinson’s disease, in view of the influence of dopaminergic regulation of beta and gamma mediators of cognitive control^13–15,19,20^.

## Methods

### Participants

Forty-four participants were recruited, comprising 21 patients with idiopathic Parkinson’s Disease (UK Brain Bank clinical diagnostic criteria) and 23 age-matched healthy controls. Four patients and two controls were excluded due to poor-compliance in-scanner with the task, or technical issues during the scanning, resulting in 17 analysed patient datasets and 21 analysed control datasets. Inclusion criteria were age 50-80 years, right-handed, and no previous history of neurological or psychiatric illness apart from Parkinson’s disease. The revised Addenbrooke’s Cognitive Assessment Scale (ACE-R), including the mini-mental state examination (MMSE), was administered to all participants. No patients had clinical presentations of dementia, and there was no difference in ACE-R scores between groups (Table 1). Patients were also assessed on the Unified Parkinson’s Disease Rating Scale^35^. Clinical assessments and experimental tasks were performed on usual dopaminergic medication, and dopaminergic dose equivalents were calculated^36^. Experimental protocols including written informed consent conformed to the guidelines of the Declaration of Helsinki and were approved by the Cambridge Research Ethics Committee (CRE code: 07/H0307/64).

**Table 1.**
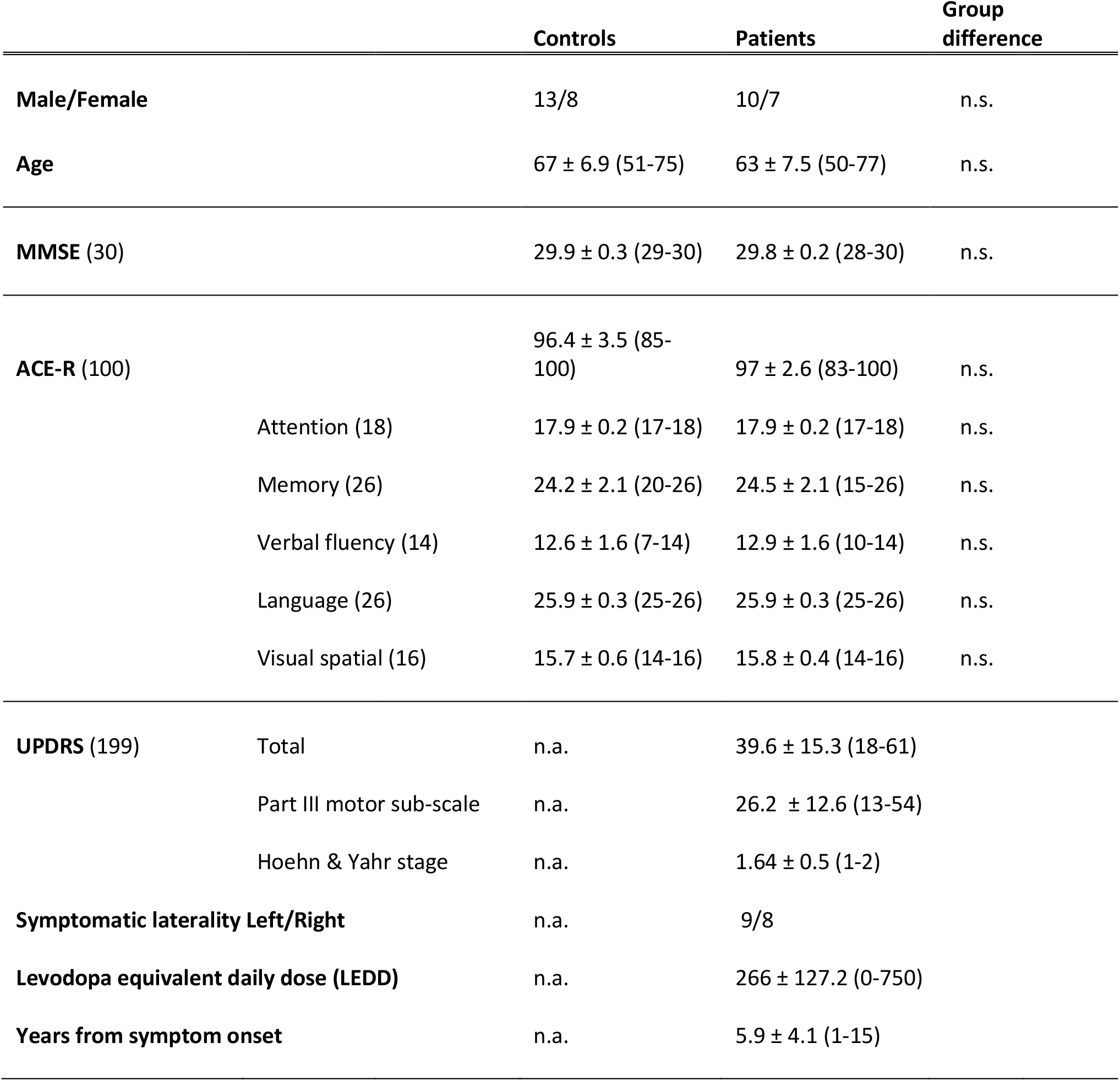
Demographic and clinical features of participants: Values shown are group means and their standard deviations (range in parentheses). MMSE: 30-point Mini Mental State Examination; ACE-R: 100-point Addenbrooke’s cognitive exam revised, divided into five subscales with maximum points in parentheses. UPDRS: Unified Parkinson’s disease rating scale. Group differences were tested using Wilcoxon rank-sum test.

|  |  | Controls | Patients | Group difference |
| --- | --- | --- | --- | --- |
| **Male/Female** |  | 13/8 | 10/7 | n.s. |
| **Age** |  | 67 ± 6.9 (51-75) | 63 ± 7.5 (50-77) | n.s. |
| **MMSE (30)** |  | 29.9 ± 0.3 (29-30) | 29.8 ± 0.2 (28-30) | n.s. |
| **ACE-R (100)** |  | 96.4 ± 3.5 (85-100) | 97 ± 2.6 (83-100) | n.s. |
|  | Attention (18) | 17.9 ± 0.2 (17-18) | 17.9 ± 0.2 (17-18) | n.s. |
|  | Memory (26) | 24.2 ± 2.1 (20-26) | 24.5 ± 2.1 (15-26) | n.s. |
|  | Verbal fluency (14) | 12.6 ± 1.6 (7-14) | 12.9 ± 1.6 (10-14) | n.s. |
|  | Language (26) | 25.9 ± 0.3 (25-26) | 25.9 ± 0.3 (25-26) | n.s. |
|  | Visual spatial (16) | 15.7 ± 0.6 (14-16) | 15.8 ± 0.4 (14-16) | n.s. |
| **UPDRS (199)** | Total | n.a. | 39.6 ± 15.3 (18-61) |  |
|  | Part III motor sub-scale | n.a. | 26.2 ± 12.6 (13-54) |  |
|  | Hoehn & Yahr stage | n.a. | 1.64 ± 0.5 (1-2) |  |
| **Symptomatic laterality Left/Right** |  | n.a. | 9/8 |  |
| **Levodopa equivalent daily dose (LEDD)** |  | n.a. | 266 ± 127.2 (0-750) |  |
| **Years from symptom onset** |  | n.a. | 5.9 ± 4.1 (1-15) |  |

### Stimuli

Stimuli were presented using Matlab (Matworks, Natiek, MA, USA) and Psychtoolbox in a quiet and dimly lit room. For training, stimuli were displayed on a CRT monitor at 60 cm distance from the observer. For the scan session, stimuli were projected on a screen at 130 cm distance (60Hz refresh rate) with equivalent pixel resolution of 0.03°. On each trial four random dot kinematograms were displayed within four circular apertures (4° diameter) positioned along a semi-circular arc (3.4° eccentricity) on a black background. 200 dots were displayed during each frame and spatially displaced to introduce apparent downward motion (6°/sec velocity). Motion coherence was manipulated by allowing only a certain proportion of dots to move downward on each frame whilst the rest were randomly reallocated. Motion coherence level was varied across trials. The 1.5 seconds long coherent motion interval was preceded and followed by intervals of zero-coherence levels lasting 1sec and 0.5sec, respectively. This was to avoid large stimulus sensory-evoked potentials elicited by abrupt stimulus onset/offset, which might mask decision processes.

### Psychophysical assessment of motion sensitivity

A practice session adopting 100% coherent stimuli allowed participants to familiarize themselves with the task. The practice session ended once participants reached 90% accuracy across all trial types. In the following psychophysical session, motion coherence was varied between trials to estimate individual motion thresholds. Eight logarithmically spaced motion coherence levels (0 0.5 0.10….0.9) were used (32 trials per level). Each training session comprised 16 blocks of 32 trials. Feedback was provided for correctness of responses as well as for too early or too late responses (100ms and 2.5s from motion coherence onset, respectively).

The psychophysical task was combined with a control match-to-sample task where occasionally (p = 0.2) after a correct choice, participants had to compare the location of a set of grey discs with the location of the previously displayed coherent stimuli. This was to ensure that participants perceived all the available options (i.e., coherent stimuli) before committing to a decision. To report a match, participants had to press any button and withhold a response otherwise. A trial was considered as correct only when both choice and matching were correct. Trials with un-matching responses were discarded and repeated within the session.

To tailor low and high levels of perceptual uncertainty to individual motion sensitivity across number of coherent stimuli, the motion-coherence dependent accuracy of each trial type (e.g., either 1 or 3 coherent stimuli) was fitted using a maximum likelihood method, with the Log-Quick function

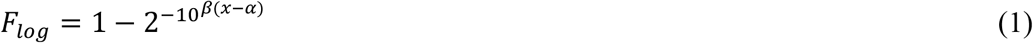

where *α* is the threshold, *β* is the slope and x is the coherence level. To obtain the proportion correct for each trial type *F*_*log*_ was scaled by

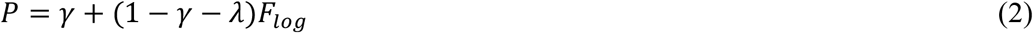

where *γ* is the guess rate and *λ* is the lapse rate controlling the lower and upper asymptote of the psychometric function, respectively.

Individual low and high perceptual uncertainty levels for each trial type were estimated as the 75th and 90th percentile of the psychometric function^15^

### Task and procedures

Participants performed a task in which they had to report a downward coherent stimulus by pressing the corresponding button^15^. The number of available coherent stimuli defined two trial types: Low action uncertainty trials, where a single coherent stimulus commanded which button to press, and high action uncertainty trials, where three coherent stimuli required the participants to press any one of the three corresponding buttons (making a “fresh choice, regardless of what you have done in previous trials”). Equal emphasis was placed on the speed and accuracy of the responses, and guessing was discouraged. Participants were instructed to fixate on a central red mark throughout the trial. Each trial started with the presentation of the fixation mark and stimuli onset ensued after a variable interval comprised between 0.5sec and 1sec. The imaging session was preceded by one training psychophysical session scheduled on a separate day (maximum 4 days between sessions). In the scan sessions, coherence levels were fixed to the individual motion thresholds corresponding to high and low levels of perceptual uncertainty, the match-to-sample task was removed, and no feedback was provided except for too early or too late responses. Levels of perceptual and action uncertainty were randomly interspersed across trials. Each session consisted of 10 blocks (total 640 trials per participant) separated by a short rest (total duration of the session ∼60 minutes).

### Voxel Based Morphometry

Following MEEG, participants underwent structural MR imaging using a 3T Siemens Tim Trio scanner with a 32-channel phased-array head coil. A T1-weighted magnetization-prepared rapid gradient-echo (MPRAGE) image was acquired with repetition time (TR)=2250ms, echo time (TE)=3.02ms, matrix=192×192, in-plane resolution of 1.25×1.25mm, 144 slices of 1.25mm thickness, inversion time=900ms and flip angle=9°. Images were first aligned to a canonical average image in MNI space, before segmentation and calculation of total intracranial volume (TIV). After segmentation, a study-specific DARTEL template was created from the patient scans and the seventeen closest age-matched controls. The remaining controls were then warped to this template. The templates were aligned to the SPM standard space and the transformation applied to all individual modulated grey-matter segments together with an 8mm FWHM Gaussian smoothing kernel.

The resulting images were entered into a full factorial general linear model with a single “group” factor of two levels, and age and TIV as covariates of no interest. This model was estimated in two steps. Firstly, a classical estimation was performed and a T-contrast constructed to compare groups. Voxels were defined as atrophic if they were statistically significant at the cluster FWE p<0.05 level. Secondly, a Bayesian estimation was performed on the same model, and a Bayesian F-contrast between patients and controls specified. The resulting Bayesian map was subjected to hypothesis testing for the null (i.e., lack of atrophy) in SPM12, resulting in a map of the posterior probability of the null at each voxel. For visualization in Figure 2, this map was thresholded for posterior probabilities for the null above 0.7 and cluster volumes of greater than 1cm^3^.

**Figure 2.**
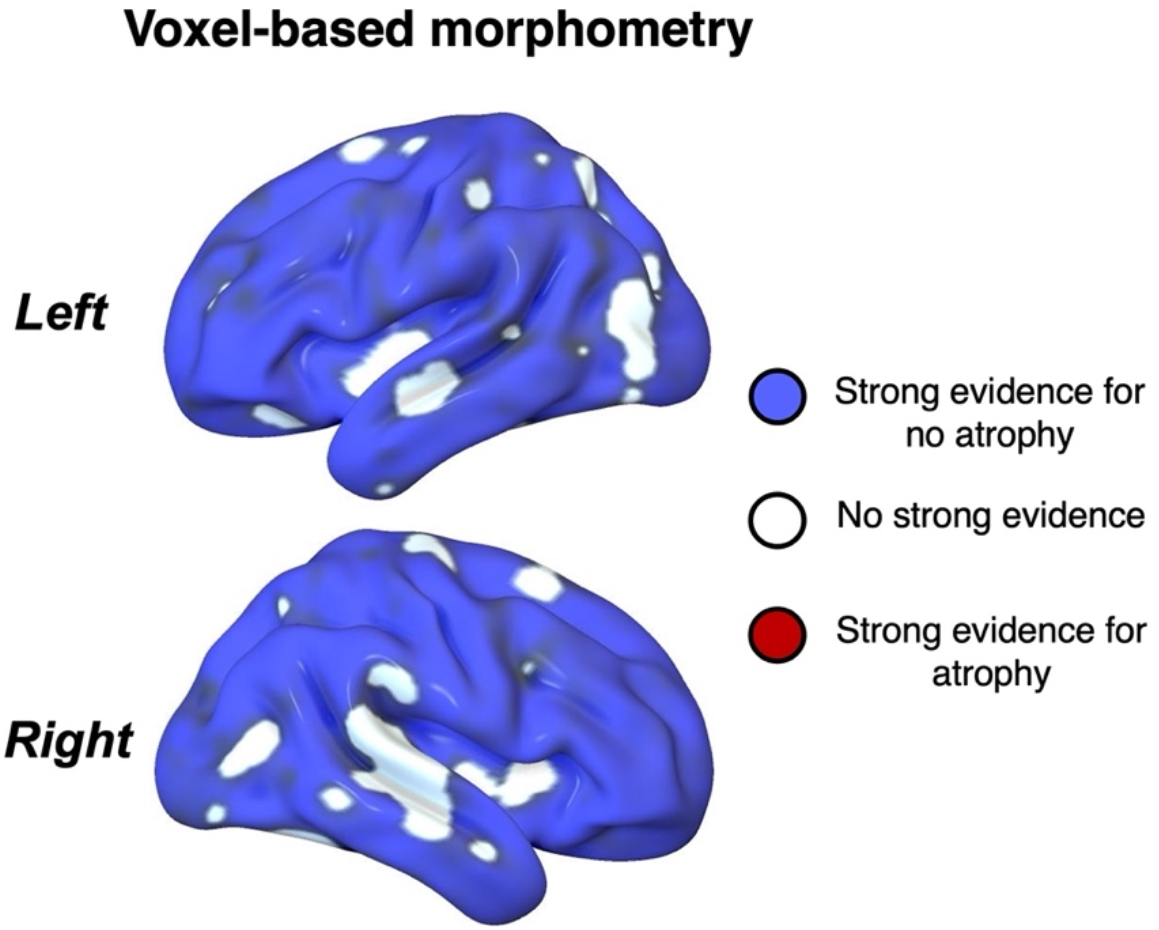
Bayesian voxel-based morphometry in the patient group. Areas in blue had strong evidence for normal cortical volume in Parkinson’s disease patients (Bayesian probability of the null > 0.7, cluster volumes > 1cm^3^). White areas had no strong evidence for or against atrophy. Red areas (not present) would indicate strong evidence for atrophy.

### MEG and EEG data acquisition and processing

An Elekta Neuromag Vectorview System (Helsinki, Finland) simultaneously acquired magnetic fields from 102 magnetometers and 204 paired planar gradiometers, and electrical potential from 70 EEG electrodes (70 Ag-AgCl scalp electrodes in an Easycap - GmbH, Herrsching, Germany - extended 10-10% system). Additional electrodes provided a nasal reference, a forehead ground, paired horizontal and vertical electro-oculography, electrocardiography and right arm electromyography. All data were recorded and digitized continuously at a sample rate of 1kHz and high-pass filtered above 0.01Hz.

Before scanning, head shape, the locations of five evenly distributed head position indicator coils, EEG electrodes location, and the position of three anatomical fiducial points (nasion and left and right pre-auricular) were recorded using a 3D digitizer (Fastrak Polhemus Inc., Colchester, VA). The initial impedance of all EEG electrodes was optimized to below 10kΩ, and if this could not be achieved in a particular channel, or if it appeared noisy to visual inspection, it was excluded from further analysis. The 3D position of the head position indicators relative to the MEG sensors was monitored throughout the scan.

These data were used by Neuromag Maxfilter 2.2 software, to perform environmental noise suppression, motion compensation, and Signal Source Separation. Subsequent analyses were performed using in-house Matlab (Mathworks) code, SPM12 (www.fil.ion.ucl.ac.uk/spm) and EEGLab (Swartz Center for Computational Neuroscience, University of California San Diego). Artifact rejection was performed through separate independent component analysis decomposition for the three sensor types. For EEG data, components temporally and spatially correlated to eye movements, blinks and cardiac activity were automatically identified with EEGLab’s toolbox ADJUST. For MEG data, components were automatically identified that were both significantly temporally correlated with electrooculography and electrocardiography data, and spatially correlated with separately acquired topographies for ocular and cardiac artifacts. A further independent component analysis identified MEG and EEG components significantly temporally correlated with tremor-related surface electromyographic activity (2-8Hz) recorded in Parkinson’s disease patients. Artifactual components were finally projected out of the dataset with a translation matrix.

The continuous artefact-corrected data were low-pass filtered (cut-off = 100Hz, Butterworth, fourth order), notch filtered between 48 and 52Hz to remove main power supply artifacts, down-sampled to 250Hz, and epoched from -1500 to 2500ms relative to motion coherence onset. EEG data were referenced to the average over electrodes. MEG and EEG data were combined before inversion into source space^37^ using the Minimum Norm algorithm as implemented by SPM12. Notably, combined MEG and EEG allows a better localization of neural sources than each technique on its own^37^. The forward model was estimated from each participant’s anatomical T1-weighted MRI image. All conditions were included in the inversion to ensure an unbiased linear mapping. The source images were spatially smoothed using an 8 mm FWHM Gaussian kernel.

### Modelling of perceptual and action decisions

To decompose behavioural performance into latent variables underpinning decisions, we fitted Linear Ballistic Accumulator models (LBA) to each participant’s reaction time data. The LBA model belongs to the broad class of accumulation-to-threshold models of decision-making but is more tractable than drift-diffusion models for n-way decisions while still remaining physiologically informative^38^. We followed the same logic of previous work^15^ and opted for a ‘unitary’ model where both perceptual and action uncertainty concur in determining a participant’s performance in a given trial. The unitary model accommodated the empirical data under the assumption of a constant non-decision time across experimental conditions, which provides theoretical evidence against the need of an extra stage of processing^39^. Accordingly, each accumulator linearly integrates the decision-evidence (or the intention) over time in favour of one action, and the decision is made when the accumulated activity reaches a threshold (see Figure 1B). In our task, possible actions correspond to a button press from one of four fingers, each modelled by independent accumulators. When three valid actions are available, three accumulators are engaged with activation starting at levels independently drawn from a uniform distribution, and increasing linearly over time with an accumulation rate *v* drawn from an independent normal distribution. A response is triggered once one accumulator wins the ‘race’ and reaches a decision bound *b*. When only one action is available, only the accumulator corresponding to the available action is engaged. Predicted reaction time is given by the duration of the accumulation process for the winning accumulator, plus a constant non-decision time *t0* representing the latency associated with stimulus encoding and motor response initiation.

To identify the combinations of free parameters that best accounted for the observed behavioural data we repeatedly fitted the LBA model with 15 unique combinations of free parameters (i.e., all possible combinations without repetition) allowed to vary across conditions. The best-fitting parameters for each model variant were used to calculate the Bayesian Information Criterion (BIC), as a measure of goodness-of fit which penalizes extra free parameters in favour of simpler models.

To adjudicate the best-fitting model variant, we adopted Bayesian random-effect analyses on the BIC values obtained by the fitting procedure^40,41^. This approach permits one to quantify the evidence for the explanatory power (i.e., frequencies) of each model across participants and groups^42^. In a first step, we assessed whether model frequencies were the same (*H*_=_) or differed (*H*_≠_) between the two groups. Under (*H*_=_) data are assumed be generated by the same model, therefore a standard random-effect analysis on the datasets pooled across groups yields the (log) evidence *p*(*Data* ∣ *H*_=_). Under *H*_≠_, group-specific datasets are assumed to be generated by different models and thus the (log) evidence *p*(*Data* ∣ *H*_≠_) is defined as the product of group-specific evidences *p*(*Controls* ∣ *H*_=_) + *p*(*Patients* ∣ *H*_=_) obtained by a separate random-effect analysis for each group. The posterior probability that the same model is valid for both groups is then given by

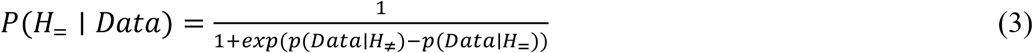

The results from the between-group model comparison confirmed that the data were generated by the same model across groups (see results). Therefore, in a second step we performed a random-effect analysis on the pooled dataset to identify the model variant prevailing in the population. The prevalence of the model was quantified as *exceedance probability*, defined as the probability that any given model is more likely than all other models^40^. Predictions of decision-related activity were generated from the winning LBA model to locate neural signatures of decisions-evidence accumulation in single-trial analyses of MEEG data^15,43^.

### Dimensionality reduction

To improve computational efficiency and reduce multiple comparisons, we reduced the dimensionality of the MEEG data by parcellating the cortical surface into a set of 96 regions of interest (ROIs) defined using the Harvard-Oxford cortical atlas (FSL, FMRIB, Oxford). The dynamic of each ROI was represented by a single time-course, obtained by extracting the principal component from the vertices lying within the given ROI^15^. The reconstructed sources within each ROI were first bandpass-filtered in either beta (13-30Hz) or gamma (31-90Hz) frequency bands. The coefficients of the principal component accounting for the majority of the variance of the vertices within each ROI, were then taken as an appropriate representation of source activity for that region. Next, to estimate the power oscillations on a single-trial basis, we extracted frequency-specific signal envelope modulations using a Hilbert transform of the source data from each reconstructed ROI (epochs from 500ms before to 1500ms after coherence onset). The Hilbert’s envelope is a convenient measure of how the power of the signal varies over time in the frequency range of interest, and thus particularly suited to capture relatively slow fluctuations associated to the instantaneous accumulation of evidence/intentions. The power estimates of individual participants were down-sampled to 100Hz and normalized by their baseline (from 400ms to 100ms before coherence onset).

### Single-trial analysis

To identify the spatio-temporal profile of decision-related accumulation over the brain we estimated the maximum lagged absolute Spearman correlation between the model predicted activity and the signal envelope in a trial-by-trial fashion^15^. Spearman correlation was chosen for its robustness to departures from normality in the power data. The lagged correlation was used to optimally split the non-decision time before and after the accumulation period to determine the time delay between the neural signal and the model predictions. The time before accumulation provides a measure of the temporal separation between sensory encoding and onset of evidence accumulation.

We estimated the largest absolute lagged correlation value for each ROI and individuals by comparing concatenated epochs and model predictions. This choice permits the measurement of accumulation lags specific to each ROI, under the assumption that they differ across brain regions for each participant. The significance of the Fisher-transformed maximum lagged correlations for each ROI was then quantified (z-score) using a non-parametric one-sample sign-test which is robust to violations of distribution symmetry. To provide a conservative estimate of significant correlations between model prediction and neural activity, we repeated the above procedure 10,000 times, each iteration using a different phase-randomized version of the original MEEG signal, to obtain a distribution of correlations under chance (null distribution). Two-tailed statistical significance was assessed by computing the proportion of absolute values from the null distribution exceeding the correlation between model predictions and the original MEEG signal. The resulting p-values were corrected for multiple comparisons (False Discovery Rate) across ROIs and frequency bands.

### Assessing the effects of uncertainty on power

To elucidate the impact of our task manipulations on power amplitude, we first averaged envelopes across trials and ROIs belonging to the dorsal path, and compared activity between low and high levels of action and perceptual uncertainty within the time window 0.1s - 1s from coherence onset, separately within each group. Significance was estimated by cluster-corrected random permutation tests (10,000 iterations, two-tailed).

Second, to explore possible moderation effects of disease on modulation of power (median over samples), we fitted a linear mixed effects model (estimated using Maximum Likelihood) where perceptual uncertainty (PU: low, high), action uncertainty (AU: low, high), and Group (control, Parkinson’s disease) were included as fixed effects. To account for individual variations in power, as well as for variation in power between brain regions, subjects and ROIs were specified as nested random effects for the model intercept (*β*_0,*i*∣*ROI*_). Two interaction terms were added to test for moderation effects of Group on PU and AU separately.

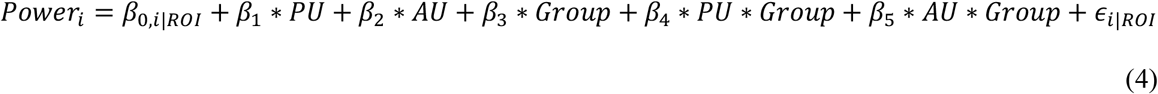

## Results

### Participant demographics

Demographic and clinical characteristics of the participants are shown in Table 1. The groups were matched for gender (Fisher’s exact test, *p*∼1.00, odds-ratio=0.88), for age (Wilcoxon rank-sum test W=120.50, *p*=0.13, two-tailed), and performed similarly on cognitive tests (ACE-R Wilcoxon rank-sum test W=161.50, *p*=0.79, two-tailed).

### Anatomical changes in Parkinson’s disease

To confirm the integrity of the network of sensory, motor and association cortices upon which our experiment relies, we used voxel-based morphometry to compare grey matter volume across the groups (Figure 2; see Table-1 for participant characteristics). We adopted classical whole brain statistical parametric mapping t-test and Bayesian null tests to assess the presence or absence of structural differences between groups. Consistent with the early stage of the disease and lack of cognitive impairment, there was evidence for lack of atrophy in most of the cortex, and no significant evidence for atrophy anywhere in cortex.

### Effects of uncertainty on behavioral performance

Individual motion coherence thresholds estimated from the training session (Figure 3) did not differ between the two groups (F_1,36_=1.48, p=0.23). Trials from the experimental session where responses were shorter than 100ms or longer than 2100ms were omitted from the behavioural and modelling analyses (controls: 15.34%, Parkinson’s disease: 20.42%). Analysis of the removed trials showed that patients did not significantly differ from controls in the number of premature responses (i.e., RT <100ms; Wilcoxon test: W=397, p=0.35, two-tailed) or omitted responses (W=345, p=0.06, two-tailed).

**Figure 3.**
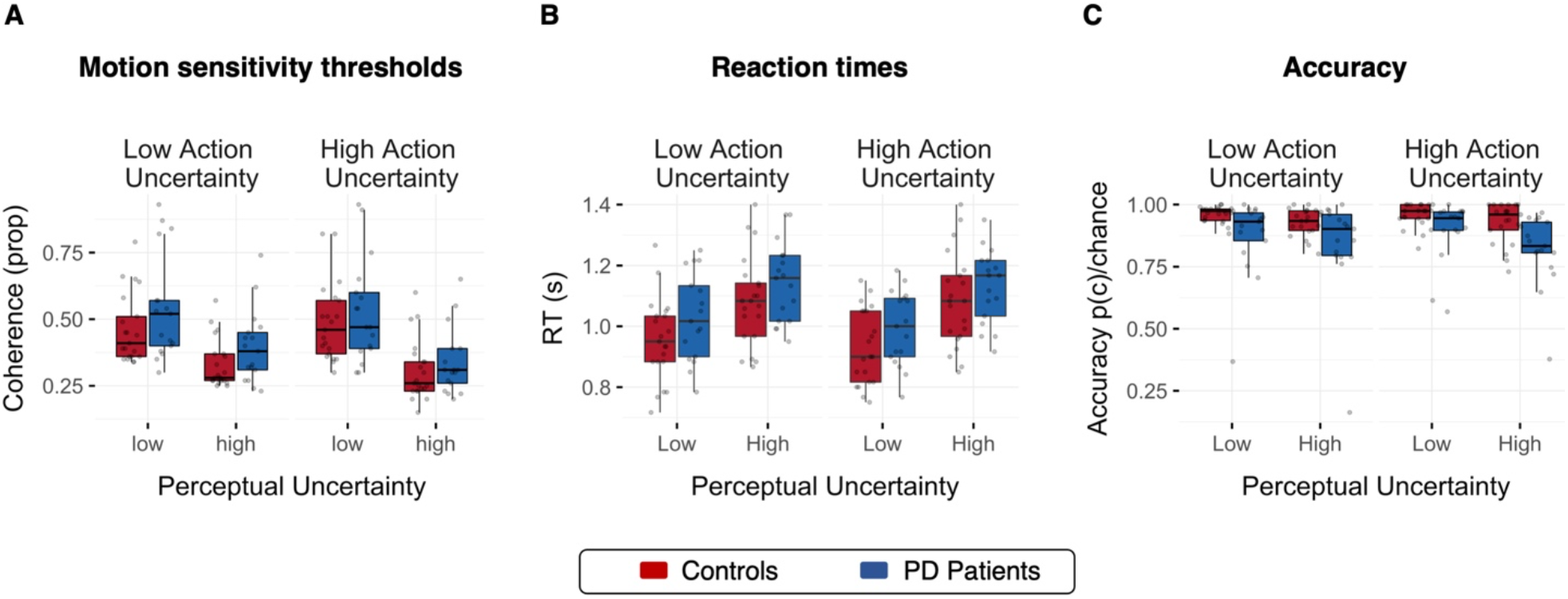
Behavioural results. **A)** Motion coherence thresholds estimated from the training session did not differ between control (red) and Parkinson’s disease (PD, blue) groups. **B)** Reaction times varied with levels of perceptual uncertainty with faster responses under low than high uncertainty. **C)** Parkinson’s disease patients were overall less accurate than controls. For both groups accuracy decreased with high perceptual uncertainty.

For the remaining trials, repeated-measures analysis of variance showed that reaction times across groups were significantly faster for the low perceptual uncertainty condition compared with the high perceptual uncertainty condition (F_1,36_=130.25, p<0.001, 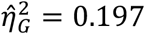, BF>100) confirming the efficacy of the estimated motion coherence thresholds. We confirmed the expected lack of a significant difference between action uncertainty levels in a n-way race^44^(F_1,36_=1.99, p=0.167, 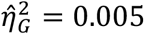, BF=0.648), and lack of interaction between action- and perceptual-uncertainty. There was no significant difference in reaction times across groups (F_1,36_=2.46, p<0.126, 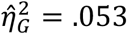, BF=1.08). Overall, Parkinson’s disease patients’ choices were less accurate than healthy controls (accuracy F_1,36_=6.77, p=0.013, 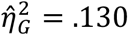, BF=4.68); For both groups, accuracy decreased with high perceptual uncertainty (F_1,36_=18.01, p<0.001, 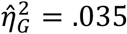, BF>100).

### Evidence accumulation in Parkinson’s disease has reduced reactivity to action uncertainty

Changes in the accumulation-rate alone (model two, Exceedance Probability=1; Figure 1B) best accounted for the effects of uncertainty on performance in both groups (*P*(*H*_=_ ∣ *Data*)=0.997, BF>100). The goodness of fit of model two (henceforth, the LBA model) was confirmed by posterior predictive checks and parameter recovery.

A mixed-design repeated-measures ANOVA on the LBA model parameters revealed slower evidence accumulation under high uncertainty levels for both action (accumulation rate: F_1,36_=190.97, MSE = 2.33, p < .001, 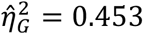, BF>100) and perceptual (accumulation rate: F_1,36_=109.22, MSE = 0.77, p < .001, 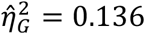, BF>100) manipulations. Overall, evidence accumulation was slower in the Parkinson’s disease group (F_1,36_=4.69, MSE = 11.41, p = 0.037, 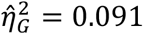, BF=2.43), with a significant interaction between group and action uncertainty (accumulation rate: F_1,36_=4.69, MSE = 11.41, p = 0.037, 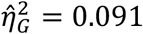, BF =26.95): controls showed larger changes in accumulation rates than patients in response to action uncertainty. Post hoc comparisons using Bonferroni corrected t-tests showed that the interaction was caused by accumulation rates significantly slower in the Parkinson’s disease group compared to controls under low action uncertainty (Low action uncertainty: *Δ* = 0.89, 95% CI[0.19, 1.59], *t*(50.10) = 2.95, *p* = 0.010; High Action uncertainty: *ΔM* = 0.30, 95% CI[−0.40, 1.00], *t*(50.10) = 1.00, *p* = 0.649).

Taken together, our results show that, regardless of group, the experimental manipulation of perceptual and action uncertainty modulated accumulation-rates in line with previous reports^2,15,43^. However, compared to healthy controls, the Parkinson’s disease group was characterized by a reduced reactivity of accumulation rates to changing uncertainty levels in the action domain (Figure 6A). Specifically, the intention to move was accumulated significantly slower than controls under low levels of action uncertainty (i.e., when a single specific action was to be taken).

### Beta power desynchronization in Parkinson’s disease has reduced reactivity to action uncertainty

The temporal evolution of the combined MEG and EEG (MEEG) power envelope from each region of interest (ROI) of the parcellated surface served as the signal for our analysis in beta (13-30Hz) and gamma (31-90Hz) bands. Confirming results from young healthy adults^15^, the LBA model predictions were inversely correlated with the MEEG oscillations in beta and gamma bands (Figure 4).

**Figure 4.**
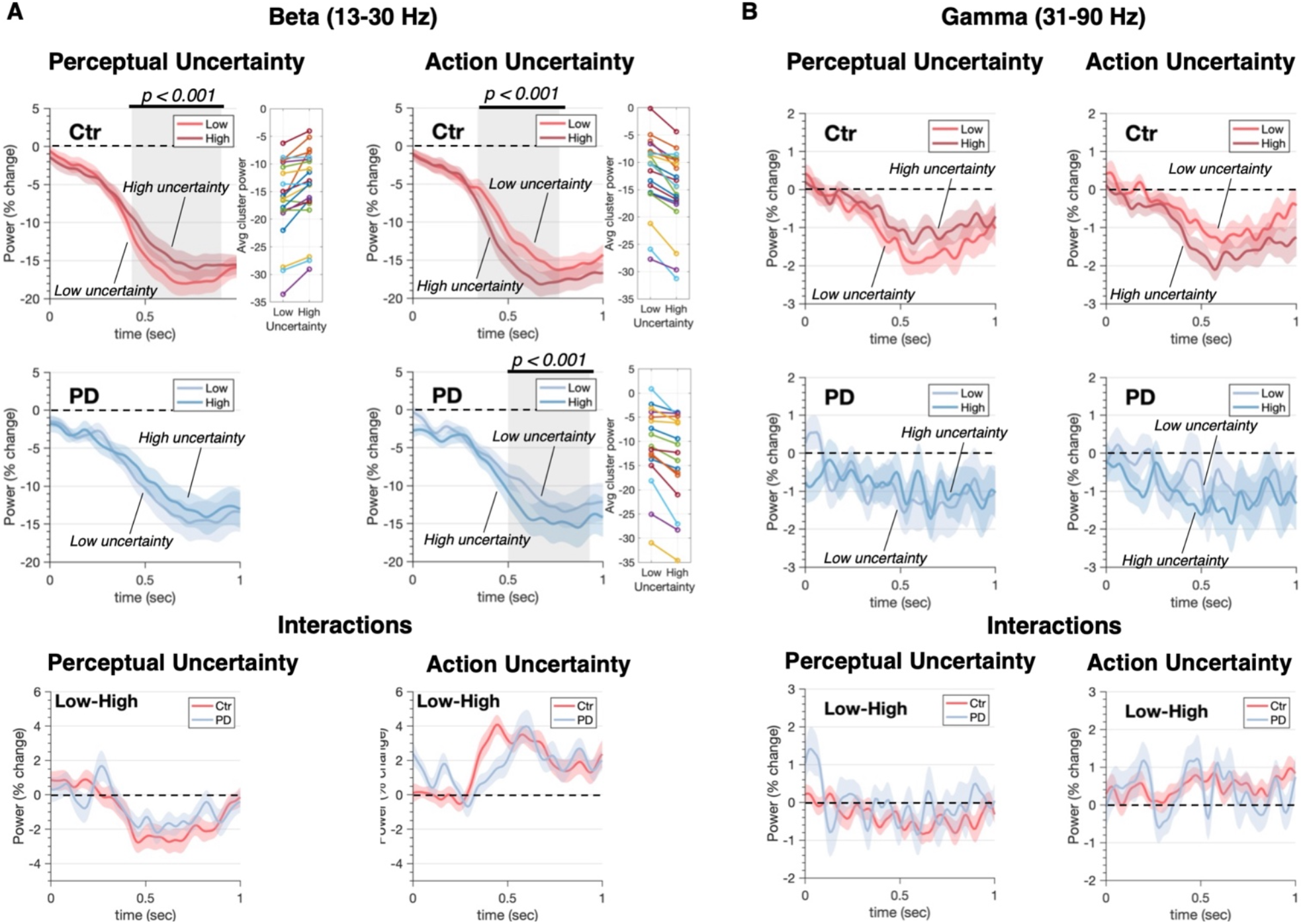
Uncertainty modulation of MEEG oscillatory activity. Power envelopes estimated for both Beta (**A**) and Gamma (**B**) bands averaged across trials and ROIs (EEG and MEG modalities fused). Top row shows power time-series for healthy controls (Ctr). Bottom row, shows the same data for Parkinson’s disease patients (PD). The onset of coherent motion was followed by desynchronization in both beta and gamma power. Desynchronization was stronger for low than high perceptual uncertainty, driven by differences in the strenght of motion signals. The pattern is reversed for action manipulations where the expected amont of total evidence accumulated scales with the number of options. The effects are significant only for beta power (with the exception of perceptual manipulation in Parkinson’s disease) and consistent across subjects: panels in the central column show individual beta power averaged within the significant cluster (participants are color-coded). Bottom panels show power changes in response to uncertainty between groups (i.e. group x uncertainty interactions). Shaded areas represent SEM, grey shaded rectangles indicate significant differences in power between low and high uncertainty levels. Significance was assessed using cluster corrected random permutations (10^3 permutations, nominal α = 0.05, Bonferroni corrected for four tests: α = 0.0125; Cohen’s d = -1.15 for perception and d = 1.88 for action comparisons in the beta range).

Specifically, after coherence onset, neural activity desynchronized in a graded fashion and peaked approximately before response suggesting a form of threshold mechanism^13,45^. For perceptual decisions, the LBA model predicts that the accumulated decision-evidence will ramp quickly with low perceptual uncertainty, and slowly with high perceptual uncertainty.

Accordingly, desynchronization of beta power-envelopes averaged across trials and ROIs was larger (p=0.0004 Bonferroni-corrected, cluster-based permutation test) for low than high perceptual uncertainty^15,45^ in controls. Such a trend was seen in the Parkinson’s disease group (p=0.088 Bonferroni-corrected). When a response is chosen between multiple options, the race underlying the selection of each alternative is characterized by a larger amount of decision-evidence summed across all the racing accumulators by the time of response^44^. Accordingly, desynchronization of beta power-envelopes averaged across trials and ROIs was larger for high than low action uncertainty in both groups (p=0.0004, Bonferroni-corrected, cluster-based permutation test). Gamma power-envelopes, showed a similar trend in the control group, but the effects were statistically insignificant. Decision-related dynamics expressed by beta desynchronization were distributed across a wide network (Figure 5A, mean sign-test z across significant ROIs, Controls: z=-5.1±0.25, p<0.0001; Patients: z=-3.99±0.58, p<0.0001; FDR-corrected) similar to previous reports^8,15^.

**Figure 5.**
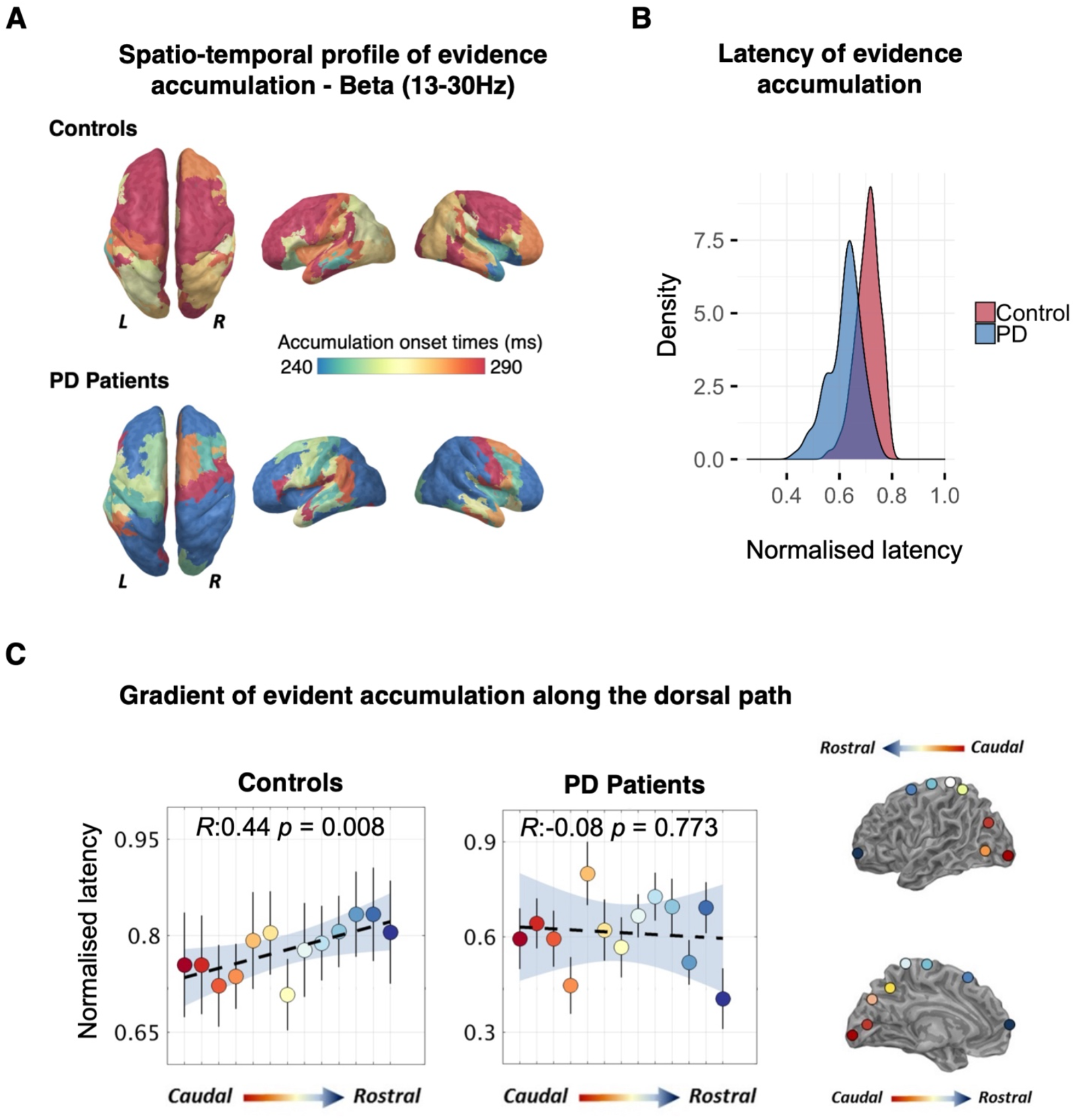
Temporal cascade of decision-evidence accumulation revealed by comparing trial-by-trial MEEG power envelopes to model’s predictions. **(A)** Latency maps showing the normalized latencies (each accumulation onset time divided by individual non-decision time) of decision-evidence accumulation mediated by beta power (13-30Hz) across anatomical regions where correlations between power-envelopes and model’s predictions survived random permutation testing. A caudo-rostral gradient is visible in the left hemisphere of healthy controls but is disrupted in Parkinson’s disease patients. **(B)** Decision-evidence accumulation in the patients group initiates slightly earlier than in controls. **(C)** Left panel: In healthy controls decision-evidence accumulation mediated by beta follows a caudo-rostral gradient along the dorsal path of the contralateral hemisphere. A linear regression best describes the gradient showing that latencies increase from visual areas up to frontal areas. In the patient group, the caudo-rostral gradient is almost inverted, with frontal regions initiating to accumulate evidence nearly in parallel with visual areas (Error bars indicate SEM, shaded area covers bootstrapped 95% regression CI). Right panel: Regions of interest (ROIs) along the dorsal path color-coded with respect to their position along the caudo-rostro axis.

Comparisons between z-transformed correlation values from each of the four levels of our manipulations in isolation confirmed that the quality of fit and results did not vary across groups and trials types (p>0.05, FDR-corrected). In the gamma band we observed a more localized mosaic of ROIs. In the control group significant ROIs included contralateral motion sensitive areas (inferior lateral occipital region), bilateral extrastriate areas and bilateral frontal regions (comprising frontal pole and superior middle gyrus), ipsilateral motor and supplementary motor area; mean across significant ROIs: sign-test z=-3.22±0.58, p=0.0031, FDR-corrected). In comparison, fewer ROIs survived statistical test in Parkinson’s disease patients (sign-test z=-2.62±0.31, p=0.0084, FDR-corrected) and none of them in the left dorsal-path, which is of primary interest for this study. Therefore, from now onwards the analyses will focus only on beta frequencies.

To explore the moderation effects of disease on beta desynchronization, we fitted a linear mixed effect model to predict power amplitude with Group (controls, patients), PU (low, high) and AU (low, high) as fixed effects, and specified nested subjects and ROIs as random factors. The model’s total explanatory power was substantial (conditional *R*^2^=0.82). The analysis confirmed the effects of perceptual (*β*=1.30, CI[1.14, 1.46], SE=0.08, p<0.0001) and action (*β*= -2.73, CI[-2.90, -2.57], SE=0.08, p<0.0001) uncertainty. A significant interaction between Group and AU indicated lower reactivity of beta power to varying levels of action uncertainty in the Parkinson’s disease group than in controls (Figure 6A; *β*=0.42, SE=0.12, CI[0.1819012 0.6673223], p<0.0001, post-hoc: Controls High-Low=-2.73, SE=0.083, t-ratio=-33, p<0.0001; Patients High-Low=-2.31, SE=0.092, t-ratio=-25, p<0.0001; Bonferroni corrected). Finally, a non-parametric Wilcoxon rank sum test confirmed exaggerated beta power (i.e., reduced desynchronization) in Parkinson’s disease compared to controls (dorsal path–both hemispheres: W=119, p=0.0416, left tailed: Controls<Patients).

**Figure 6.**
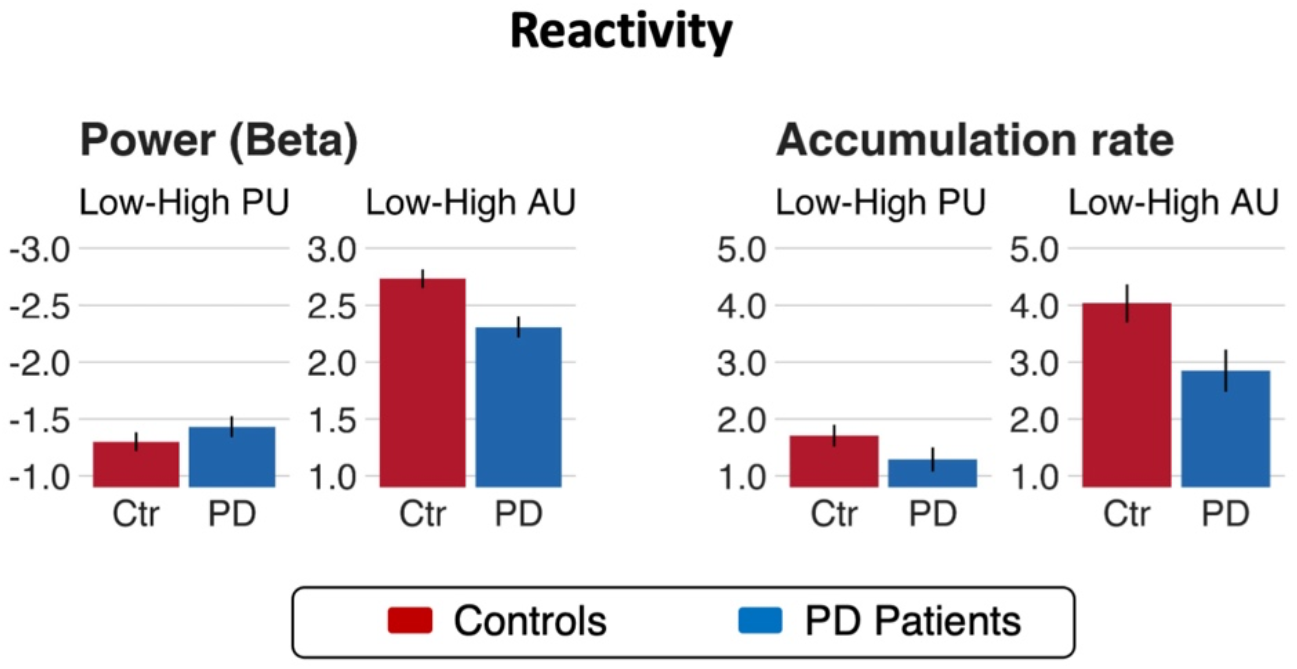
Parkinson’s disease is associated with reduced flexibility and disrupted gradient of evidence accumulation. Lower reactivity of beta power (left panel) and accumulation-rate (right panel) to varying levels of action uncertainty in the Parkinson’s disease group than in controls.

### Evidence accumulation cascade is dysregulated in Parkinson’s disease

To examine the decision-evidence accumulation over space and time, the onset of evidence accumulation across ROIs was identified by optimizing the split of the non-decision time before and after the accumulation period using Spearman correlation to the single-trial MEEG power envelope (Figure 5A, see Methods and Materials for details). By tracing the spectrally resolved temporal evolution of decision onset through the visuo-motor hierarchy we found that decision-evidence accumulation emerges with distinct spatio-temporal profiles between healthy controls and Parkinson’s disease patients (Figure 5A-C). In the control group we replicated the results from Tomassini et al.^15^. Specifically, we show that in the contralateral (i.e., left) hemisphere, the beta-mediated evidence accumulation unfolds in a bottom-up cascade proceeding from caudal sensorial regions to rostral executive areas (Figure 5A, C).

In people with Parkinson’s disease, accumulation in the beta-frequency begins *earlier* than in controls (∼240ms from coherence onset; Wilcoxon rank-sum: W=7977, p<0.001). Crucially, the caudo-rostral beta gradient of evidence accumulation is abolished, with frontal regions beginning to accumulate evidence in parallel with caudal visual areas.

The difference between groups is shown in Figure 5. We fitted a regression model to the mean latencies of ROIs located along the dorsal path for visuomotor decisions^46,47^(Figure 5C). For controls (Figure 5C left top-bottom panels) there is a gradient from caudal to rostral regions (R^2^=0.44, p=0.008), which contrasts the lack of gradient in Parkinson’s disease patients (R^2^= -0.08, p=0.733). This indicates early decision processes begin in the frontal pole and the middle frontal gyrus, preceding (∼150ms) onsets in the occipital pole.

## Discussion

The principal result of this study is that Parkinson’s disease alters the spatiotemporal cascade of decisions involved in the transformation from visual stimuli to motor response. Whereas healthy adults manifest a caudal-to-rostral gradient in the latency to onset of evidence accumulation in the beta-frequency range, this gradient is lost in people with Parkinson’s disease (Figure 5A).

Moreover, the accumulation of evidence in the beta range is inflexible in people with Parkinson’s disease, with loss of the modulation according to the perceptual or action uncertainty^2,34,48^(Figure 6). Short sensory encoding (i.e., the pre-decision component of the non-decision time) and early accumulation of evidence in the beta-range are not confined to classical “visual processing” regions, but are observed throughout the dorsal stream. Indeed, the most striking difference in Figure 5A is seen over frontal cortical regions, consistent with early proactive rather than later reactive escalation of beta-power in frontal sources of top-down influence on the visuomotor decision process. Behaviourally, this was reflected in more errors and slower response times in Parkinson’s disease (Figure 3), despite normal cortical volume (Figure 2).

The use of accumulation-to-threshold modelling enables the identification of anatomical and frequency-specific correlates of the latent cognitive processes underpinning sensorimotor decisions. The modelling of behavioural performance confirmed that increasing levels of perceptual and action uncertainty normally affect decisions by slowing the speed of evidence accumulation in both groups^2,15,48^. The impact of action uncertainty was different in people with Parkinson’s disease. We refer to the accumulated evidence of action as ‘intention’, to indicate its association with the response rather than stimulus. The accumulation of such action intentions was slower and inflexible in patients compared to controls^2,21^ (Figure 6A), in keeping with other markers of cognitive inflexibility in Parkinson’s disease^3,49,50^.

In both groups, the within-trial accumulation of evidence was correlated with changes in beta and gamma power. Beta-desynchronization has previously been shown to scale with uncertainty^45^. Here, the temporal profile displays a signature accumulation of decision-evidence over time to a consistent bound that is reached shortly before each movement^13,51,52^. However, beta desynchronization was impaired by Parkinson’s disease, not only in association with making the response^53–56^ but also in the loss of sensitivity to uncertainty. Note that in our study, cortical beta-desynchronization was recorded by MEEG, not the subthalamic beta desynchronization that is also abnormal in Parkinson’s disease^57,58^. Changes in desynchronization are related to accuracy and latency^59,60^. Faster beta-mediated flow of decision-evidence through a visuo-motor processing hierarchy is associated with faster and more accurate decisions^15^. During sensory discrimination by non-human primates^61^, beta desynchronization is greater for accurate trials.

Beta desynchronization may represent a general control process not just a determinant of movement. For example, lateralized beta desynchronization correlates with movement preparation, as well as the state of decision-evidence and the updating of a motor plan as decision evolves^8,13,45,52,62^.

In controls, the effect of uncertainty on the evolution of gamma power was qualitatively similar to the effect on beta power, but not significant when permutation corrected for multiple comparisons (Figure 4B), likely due to the lower signal-to-noise ratio of magnetoencephalography at high frequencies. In people with Parkinson’s disease, the effect of uncertainty on gamma power was not seen, but we acknowledge that even the group average data are clearly very noisy. Gamma activity has been proposed to bring neural circuits into a state of readiness for the visuomotor processing^63^, routing task-relevant information through the integrator units. Such a role may be a specific manifestation of the broader role of gamma oscillations in feed-forward signalling through cortical networks^64,65^. We attribute the desynchronization of gamma to the loss of sensory information content when the random dot kinematogram switches to coherence: although the direction of coherence is new information, the pre-coherence motion of all 200 dots has more un-predicted information over the sum of all stimuli.

In people with Parkinson’s disease, the change in beta power and loss of beta-reactivity are common signatures of pathology in the basal ganglia and its effect on frontoparietal network function^19^, and on cognitive or motor processes^20,66^. Our patients were on their usual dopaminergic medication, that is usually clinically optimised according to motor function, rather than cognition in the absence of clinical cognitive impairment. Deficits in dopamine-dependent pathways may therefore still cause the aberrant generation of oscillatory activity in the beta range during cognitive operations, including decisions for the selection of responses^19,67^. Our model of evidence accumulation posits that evolving perceptual decisions inform action selection accumulators, enabling a parallel deliberation of available options and motor responses^7–10^.

Posterior regions in health start accumulating evidence first (as soon as sensory information is encoded), but in Parkinson’s disease patients, integration of in occipital and frontal cortex begins near simultaneously. This suggests that the accumulated evidence (or intention) for action selection is no longer dependent on the evolving perceptual decision, but may instead draw preferentially on prior expectations or perseverated responses.

The normal cascade establishes a compromise between the speed of parallel processes and the accuracy of robust serial decisions. A consequence of the changes in Parkinson’s disease we observe is therefore to improve speed of responding at the potential expense of optimal action selection. The patient responses are correspondingly fast but inaccurate (Figure 3B-C).

Conversely, a delay is typically associated with improved accuracy^68–71^. Since the aim of sequentially sampling noisy evidence over time is to limit the impact of noise on deliberation, one implication of insufficient sampling before decision onset on downstream accumulators is low quality and less precise (i.e., noisier) information entering the decision process^51^. The optimal updating of a decision in light of new evidence can also be described as the balanced influence of bottom-up new evidence and top-down prior evidence^8,72^ (Figure 6B). Noisy sensory information is effectively down-weighted in favour of more precise (stronger) predictions encoded by top-down priors. Recent work on perceptual decision-making^73^ and visuomotor control^74^ suggests shift towards top-down control in people with Parkinson’s disease, which we speculate to arise from the earlier than normal beta power accumulation in prefrontal cortex. Beta oscillations may be a neurophysiological correlate of the estimate of bottom-up uncertainty^74–76^ tracking the trial-by-trial weight of evidence for making decisions^8^, and the balance between bottom-up evidence vs top-down control during decision-making^8,74,77–79^. While slower response is typically associated with improved accuracy^68–71^, patients may be partially compensating for slower action selection and inflexible behaviour by reducing the time allocated to the accumulation of sensory evidence, sacrificing accuracy to keep behavioural reactions within an ecologically balanced range.

There are limitations to our study. We rely on clinical diagnosis, without evidence of neuropathology in our patient cohort. Our sample size was modest, although in keeping with medium to large effect sizes in previous work on motor control and action selection in Parkinson’s disease, and large in the context of the task-based magnetoencephalography literature. Further, our participants were on their usual medication and we cannot confirm the dopaminergic basis of the effects we observe, as opposed to other anatomical and neurochemical consequences of the disease. Nonetheless, we did confirm that our participants did not have dementia or marked cognitive impairment, or cortical atrophy. The MEEG method does not detect subcortical signals, but is restricted to cortical neurophysiology. The lack of atrophy, together with the condition-specific and frequency-specific effects we observed, makes it less unlikely that a non-specific cognitive impairment is the cause of the observed abnormalities. Nonetheless, we are agnostic as to whether the observed neurophysiological changes are a direct result of cortical pathology of indirect consequence of subcortical degeneration in cortico-striato-thalamo-cortical circuits and their dopaminergic innervation.

In conclusion, we have demonstrated the integration of cortical physiological recordings with linear ballistic accumulator models of decisions, based on sensory evidence and motor intentions. The normal cascade of temporally overlapping decisions, with a rostro-causal gradient of latency of beta-mediated accumulation, is absent in Parkinson’s disease. This is accompanied by insensitivity of the beta power to uncertainty, representing the failure to modify decision processes in the face of uncertainty which is ordinarily required to optimise behavioural decisions.

## Data Availability

All data produced in the present study are available upon reasonable request to the authors. Data and code to reproduce figures will be made available online upon acceptance of the manuscript.

## References

1. Weintraub D, Koester J, Potenza MN, et al. Impulse Control Disorders in Parkinson Disease. Arch Neurol. 2010;67(5):589–595. doi:10.1001/archneurol.2010.65

2. Zhang J, Rittman T, Nombela C, et al. Different decision deficits impair response inhibition in progressive supranuclear palsy and Parkinson’s disease. Brain. 2016;139(1):161–173. doi:10.1093/brain/awv331

3. Robbins TW, Cools R. Cognitive deficits in Parkinson’s disease: A cognitive neuroscience perspective. Mov Disord. 2014;29(5):597–607. doi:10.1002/mds.25853

4. Napier TC, Corvol JC, Grace AA, et al. Linking neuroscience with modern concepts of impulse control disorders in Parkinson’s disease. Mov Disord. 2015;30(2):141–149. doi:10.1002/MDS.26068

5. Lawson RA, Yarnall AJ, Duncan GW, et al. Cognitive decline and quality of life in incident Parkinson’s disease: The role of attention. Parkinsonism Relat Disord. 2016;27:47–53. doi:10.1016/J.PARKRELDIS.2016.04.009

6. McLelland J. On the Time Relations of Mental Processes: An Examination of Systems of Processes in Cascade.; 1979.

7. Heekeren HR, Marrett S, Ungerleider LG. The neural systems that mediate human perceptual decision making. Nat Rev Neurosci. 2008;9(6):467–479. doi:10.1038/nrn2374

8. Gould IC, Nobre a. C, Wyart V, Rushworth MFS. Effects of Decision Variables and Intraparietal Stimulation on Sensorimotor Oscillatory Activity in the Human Brain. J Neurosci. 2012;32(40):13805–13818. doi:10.1523/JNEUROSCI.2200-12.2012

9. Lange FP De, Rahnev D a, Donner TH, Lau H. Prestimulus Oscillatory Activity over Motor Cortex Reflects Perceptual Expectations. J Neurosci. 2013;33(4):1400–1410. doi:10.1523/JNEUROSCI.1094-12.2013

10. Siegel M, Buschman TJ, Miller EK. Cortical information flow during flexible sensorimotor decisions. Science (80-). 2015;348(6241):1352–1355. doi:10.1126/science.aab0551

11. Zhang J, Hughes LE, Rowe JB. Selection and inhibition mechanisms for human voluntary action decisions. Neuroimage. 2012;63(1):392–402. doi:10.1016/j.neuroimage.2012.06.058

12. Fries P. Rhythms for Cognition: Communication through Coherence. Neuron. 2015;88(1):220–235. doi:10.1016/j.neuron.2015.09.034

13. Donner TH, Siegel M, Fries P, Engel AK. Buildup of Choice-Predictive Activity in Human Motor Cortex during Perceptual Decision Making. Curr Biol. 2009;19(18):1581–1585. doi:10.1016/j.cub.2009.07.066

14. Polanía R, Krajbich I, Grueschow M, Ruff CC. Neural Oscillations and Synchronization Differentially Support Evidence Accumulation in Perceptual and Value-Based Decision Making. Neuron. 2014;82:709–720. doi:10.1016/j.neuron.2014.03.014

15. Tomassini A, Price D, Zhang J, Rowe J. On the evolution of neural decisions from uncertain visual input to uncertain actions. Published online 2020:1–46. doi:10.1101/803049

16. Salenius S, Avikainen S, Kaakkola S, Hari R, Brown P. Defective cortical drive to muscle in Parkinson’s disease and its improvement with levodopa. Brain. 2002;125(Pt 3):491–500. doi:10.1093/BRAIN/AWF042

17. Stoffers D, Bosboom JLW, Wolters EC, Stam CJ, Berendse HW. Dopaminergic modulation of cortico-cortical functional connectivity in Parkinson’s disease: an MEG study. Exp Neurol. 2008;213(1):191–195. doi:10.1016/J.EXPNEUROL.2008.05.021

18. Schnitzler A, Gross J. Normal and pathological oscillatory communication in the brain. Nat Rev Neurosci. 2005;6(4):285–296. doi:10.1038/nrn1650

19. West TO, Berthouze L, Halliday DM, et al. Propagation of beta/gamma rhythms in the cortico-basal ganglia circuits of the parkinsonian rat. J Neurophysiol. 2018;119(5):1608–1628. doi:10.1152/jn.00629.2017

20. Oswal A, Litvak V, Sauleau P, Brown P. Beta reactivity, prospective facilitation of executive processing, and its dependence on dopaminergic therapy in Parkinson’s disease. J Neurosci. 2012;32(29):9909–9916. doi:10.1523/JNEUROSCI.0275-12.2012

21. O’Callaghan C, Hall JM, Tomassini A, et al. Visual Hallucinations Are Characterized by Impaired Sensory Evidence Accumulation: Insights From Hierarchical Drift Diffusion Modeling in Parkinson’s Disease. Biol Psychiatry Cogn Neurosci Neuroimaging. 2017;2(8):680–688. doi:10.1016/j.bpsc.2017.04.007

22. Tomassini A, Pollak TA, Edwards MJ, Bestmann S. Learning from the past and expecting the future in Parkinsonism: Dopaminergic influence on predictions about the timing of future events. Neuropsychologia. 2019;b127. doi:10.1016/j.neuropsychologia.2019.02.003

23. Murray EA, Rudebeck PH. Specializations for reward-guided decision-making in the primate ventral prefrontal cortex. Nat Rev Neurosci. Published online 2018:1. doi:10.1038/s41583-018-0013-4

24. Phillips HN, Cope TE, Hughes LE, Zhang J, Rowe JB. Monitoring the past and choosing the future: the prefrontal cortical influences on voluntary action. Sci Rep. 2018;8(1):7247. doi:10.1038/s41598-018-25127-y

25. Mulder MJ, van Maanen L, Forstmann BU. Perceptual decision neurosciences - a model-based review. Neuroscience. 2014;277:872–884. doi:10.1016/j.neuroscience.2014.07.031

26. Brown SD, Heathcote A. The simplest complete model of choice response time: Linear ballistic accumulation. Cogn Psychol. 2008;57:153–178. doi:10.1016/j.cogpsych.2007.12.002

27. Kiani R, Hanks TD, Shadlen MN. Bounded integration in parietal cortex underlies decisions even when viewing duration is dictated by the environment. J Neurosci. 2008;28(12):3017–3029. doi:10.1523/JNEUROSCI.4761-07.2008

28. Resulaj A, Kiani R, Wolpert DM, Shadlen MN. Changes of mind in decision-making. Nature. 2009;461(7261):263–266. doi:10.1038/nature08275

29. Roitman JD, Shadlen MN. Response of neurons in the lateral intraparietal area during a combined visual discrimination reaction time task. J Neurosci. 2002;22(21):9475–9489. doi:10.1016/S0377-2217(02)00363-6

30. Teichert T, Grinband J, Ferrera V. The importance of decision onset. J Neurophysiol. 2016;115(2):643–661. doi:10.1152/jn.00274.2015

31. Laming D. Choice reaction performance following an error. Acta Psychol (Amst). 1979;43(3):199–224. doi:10.1016/0001-6918(79)90026-X

32. O’Callaghan C, Lewis SJG. Cognition in Parkinson’s Disease. Int Rev Neurobiol. 2017;133:557–583. doi:10.1016/bs.irn.2017.05.002

33. Jiaxiang Zhang, Timothy Rittman, Cristina Nombela, Alessandro Fois, Ian Coyle-Gilchrist, Roger A. Barker LEH and JBR. Different decision deficits impair response inhibition in progressive supranuclear palsy and Parkinson’s disease. Brain. Published online 2015. doi:10.3102/0002831212437854

34. Zhang J, Nombela C, Wolpe N, Barker RA, Rowe JB. Time on timing: Dissociating premature responding from interval sensitivity in Parkinson’s disease. Mov Disord. 2016;31(8):1163–1172. doi:10.1002/mds.26631

35. Goetz CG, Tilley BC, Shaftman SR, et al. Movement Disorder Society-sponsored revision of the Unified Parkinson’s Disease Rating Scale (MDS-UPDRS): scale presentation and clinimetric testing results. Mov Disord. 2008;23(15):2129–2170. doi:10.1002/MDS.22340

36. Tomlinson CL, Stowe R, Patel S, Rick C, Gray R, Clarke CE. Systematic review of levodopa dose equivalency reporting in Parkinson’s disease. Mov Disord. 2010;25(15):2649–2653. doi:10.1002/mds.23429

37. Henson RN, Mouchlianitis E, Friston KJ. MEG and EEG data fusion : Simultaneous localisation of face-evoked responses. Neuroimage. 2010;47(2):581–589. doi:10.1016/j.neuroimage.2009.04.063.MEG

38. Cassey P, Heathcote A, Brown SD. Brain and Behavior in Decision-Making. PLoS Comput Biol. 2014;10(7). doi:10.1371/journal.pcbi.1003700

39. Ratcliff R, McKoon G. The Diffusion Decision Model: Theory and Data for Two-Choice Decision Tasks. Neural Comput. 2008;20(4):873–922. doi:10.1162/neco.2008.12-06-420

40. Stephan KE, Penny WD, Daunizeau J, Moran RJ, Friston KJ. Bayesian model selection for group studies. Neuroimage. 2009;46(4):1004–1017. doi:10.1016/j.neuroimage.2009.03.025

41. Tomassini A, Ruge D, Galea JM, Penny W, Bestmann S. The Role of Dopamine in Temporal Uncertainty. J Cogn Neurosci. Published online September 24, 2015:1–15. doi:10.1162/jocn_a_00880

42. Daunizeau J, Adam V, Rigoux L. VBA: A Probabilistic Treatment of Nonlinear Models for Neurobiological and Behavioural Data. Prlic A, ed. PLoS Comput Biol. 2014;10(1):e1003441. doi:10.1371/journal.pcbi.1003441

43. Rowe JB, Hughes LE, Barker R a, Owen a M. Dynamic causal modelling of effective connectivity from fMRI: are results reproducible and sensitive to Parkinson’s disease and its treatment? Neuroimage. 2010;52(3):1015–1026. doi:10.1016/j.neuroimage.2009.12.080

44. Rowe JB, Hughes L, Nimmo-Smith I. Action selection: a race model for selected and non-selected actions distinguishes the contribution of premotor and prefrontal areas. Neuroimage. 2010;51(2):888–896. doi:10.1016/j.neuroimage.2010.02.045

45. Kubanek J, Snyder LH, Brunton BW, Brody CD, Schalk G. A low-frequency oscillatory neural signal in humans encodes a developing decision variable. Neuroimage. 2013;83:795–808. doi:10.1016/j.neuroimage.2013.06.085

46. Donner TH, Siegel M, Oostenveld R, Fries P, Bauer M, Engel AK. Population activity in the human dorsal pathway predicts the accuracy of visual motion detection. J Neurophysiol. 2007;98(1):345–359. doi:10.1152/jn.01141.2006

47. Kopell N, Ermentrout GB, Whittington MA, Traub RD. Gamma rhythms and beta rhythms have different synchronization properties. Proc Natl Acad Sci. 2000;97(4):1867–1872. doi:10.1073/pnas.97.4.1867

48. Frank MJ, Samanta J, Moustafa A a, Sherman SJ. Hold your horses: impulsivity, deep brain stimulation, and medication in parkinsonism. Science. 2007;318(2007):1309–1312. doi:10.1126/science.1146157

49. Cools R, Altamirano L, D’Esposito M. Reversal learning in Parkinson’s disease depends on medication status and outcome valence. Neuropsychologia. 2006;44(10):1663–1673. doi:10.1016/J.NEUROPSYCHOLOGIA.2006.03.030

50. Kehagia AA, Cools R, Barker RA, Robbins TW. Switching between abstract rules reflects disease severity but not dopaminergic status in Parkinson’s disease. Neuropsychologia. 2009;47(4):1117–1127. doi:10.1016/J.NEUROPSYCHOLOGIA.2009.01.002

51. Gold JI, Shadlen MN. The neural basis of decision making. Annu Rev Neurosci. 2007;30:535–574. doi:10.1146/annurev.neuro.29.051605.113038

52. Wyart V, de Gardelle V, Scholl J, Summerfield C. Rhythmic Fluctuations in Evidence Accumulation during Decision Making in the Human Brain. Neuron. 2012;76(4):847–858. doi:10.1016/j.neuron.2012.09.015

53. Wu HM, Hsiao FJ, Chen RS, et al. Attenuated NoGo-related beta desynchronisation and synchronisation in Parkinson’s disease revealed by magnetoencephalographic recording. Sci Rep. 2019;9(1). doi:10.1038/S41598-019-43762-X

54. te Woerd ES, Oostenveld R, de Lange FP, Praamstra P. A shift from prospective to reactive modulation of beta-band oscillations in Parkinson’s disease. Neuroimage. 2014;100:507–519. doi:10.1016/J.NEUROIMAGE.2014.06.039

55. Heinrichs-Graham E, Wilson TW, Santamaria PM, et al. Neuromagnetic evidence of abnormal movement-related beta desynchronization in Parkinson’s disease. Cereb Cortex. 2014;24(10):2669–2678. doi:10.1093/CERCOR/BHT121

56. Brown P, Marsden D. Bradykinesia and Impairment of EEG Desynchronization in Parkinson’s Disease. Published online 1999. doi:10.1002/1531-8257

57. Kühn AA, Doyle L, Pogosyan A, et al. Modulation of beta oscillations in the subthalamic area during motor imagery in Parkinson’s disease. Brain. 2006;129(Pt 3):695–706. doi:10.1093/BRAIN/AWH715

58. Eisinger RS, Cagle JN, Opri E, et al. Parkinsonian Beta Dynamics during Rest and Movement in the Dorsal Pallidum and Subthalamic Nucleus. J Neurosci. 2020;40(14):2859–2867. doi:10.1523/JNEUROSCI.2113-19.2020

59. Frey JN, Ruhnau P, Weisz N. Not so different after all: The same oscillatory processes support different types of attention. Brain Res. 2015;1626:183–197. doi:10.1016/j.brainres.2015.02.017

60. Hanslmayr S, Staudigl T, Fellner MC. Oscillatory power decreases and long-term memory: the information via desynchronization hypothesis. Front Hum Neurosci. 2012;6(April):1–12. doi:10.3389/fnhum.2012.00074

61. Haegens S, Nacher V, Hernandez A, Luna R, Jensen O, Romo R. Beta oscillations in the monkey sensorimotor network reflect somatosensory decision making. Proc Natl Acad Sci. 2011;108(26):10708–10713. doi:10.1073/pnas.1107297108

62. Engel AK, Fries P. Beta-band oscillations-signalling the status quo? Curr Opin Neurobiol. 2010;20(2):156–165. doi:10.1016/j.conb.2010.02.015

63. Olufsen MS, Whittington MA, Camperi M, Kopell N. New roles for the gamma rhythm: Population tuning and preprocessing for the beta rhythm. J Comput Neurosci. 2003;14(1):33–54. doi:10.1023/A:1021124317706

64. Bastos AM, Lundqvist M, Waite AS, Kopell N, Miller EK. Layer and rhythm specificity for predictive routing. Proc Natl Acad Sci U S A. 2020;117(49):31459–31469. doi:10.1073/PNAS.2014868117

65. Vezoli J, Vinck M, Bosman CA, et al. Brain rhythms define distinct interaction networks with differential dependence on anatomy. Neuron. 2021;109(23):3862-3878.e5. doi:10.1016/J.NEURON.2021.09.052

66. Boon LI, Geraedts VJ, Hillebrand A, et al. A systematic review of MEG-based studies in Parkinson’s disease: The motor system and beyond. Hum Brain Mapp. 2019;40(9):2827–2848. doi:10.1002/hbm.24562

67. Rowe JB, Hughes L, Ghosh BCP, et al. Parkinson ’ s disease and dopaminergic therapyçdifferential effects on movement, reward and cognition. Published online 2008:2094–2105. doi:10.1093/brain/awn112

68. Pouget P, Logan GD, Palmeri TJ, Boucher L, Paré M, Schall JD. Neural basis of adaptive response time adjustment during saccade countermanding. J Neurosci. 2011;31(35):12604–12612. doi:10.1523/JNEUROSCI.1868-11.2011

69. Purcell BA, Schall JD, Logan GD, Palmeri TJ. From salience to saccades: multiple-alternative gated stochastic accumulator model of visual search. J Neurosci. 2012;32(10):3433–3446. doi:10.1523/JNEUROSCI.4622-11.2012

70. Ratcliff R, Frank MJ. Reinforcement-based decision making in corticostriatal circuits: mutual constraints by neurocomputational and diffusion models. Neural Comput. 2012;24(5):1186–1229. doi:10.1162/NECO_a_00270

71. Schmitz F, Voss A. Decomposing task-switching costs with the diffusion model. J Exp Psychol Hum Percept Perform. 2012;38(1):222–250. doi:10.1037/a0026003

72. Friston KJ. A theory of cortical responses. Philos Trans R Soc Lond B Biol Sci. 2005;360:815–836.

73. Vilares I, Kording KP. Dopaminergic medication increases reliance on current information in Parkinson’s disease. Nat Hum Behav. 2017;1(8):0129. doi:10.1038/s41562-017-0129

74. Palmer CE, Auksztulewicz R, Ondobaka S, Kilner JM. Sensorimotor beta power reflects the precision-weighting afforded to sensory prediction errors. Neuroimage. 2019;200(February):59–71. doi:10.1016/j.neuroimage.2019.06.034

75. Palmer C, Zapparoli L, Kilner JM. A New Framework to Explain Sensorimotor Beta Oscillations. Trends Cogn Sci. 2016;20(5):321–323. doi:10.1016/j.tics.2016.03.007

76. Sedley W, Gander PE, Kumar S, et al. Neural signatures of perceptual inference. Elife. 2016;5(MARCH 2016). doi:10.7554/ELIFE.11476

77. De Lange FP, Jensen O, Dehaene S. Accumulation of evidence during sequential decision making: The importance of top-down factors. J Neurosci. 2010;30(2):731–738. doi:10.1523/JNEUROSCI.4080-09.2010

78. Arnal LH, Giraud AL. Cortical oscillations and sensory predictions. Trends Cogn Sci. 2012;16(7):390–398. doi:10.1016/J.TICS.2012.05.003

79. Cope TE, Sohoglu E, Sedley W, et al. Evidence for causal top-down frontal contributions to predictive processes in speech perception. Nat Commun. 2017;8(1). doi:10.1038/S41467-017-01958-7

